# Death profiling of hospitalized patients with COVID-19: Experience from a specialized hospital in Bangladesh

**DOI:** 10.1101/2021.07.04.21259979

**Authors:** Md. Shahed-Morshed, Abdullah Al Mosabbir, Mohammad Sorowar Hossain

**Author notes:** **Corresponding author:** Mohammad Sorowar Hossain; PhD, Department of Emerging and Neglected Diseases, Biomedical Research Foundation, Dhaka, Bangladesh.

## Abstract

**Background:** The information on characteristics and causes of mortality in deceased patients with coronavirus disease 2019 (COVID-19) is scarce in the literature. This study aimed to document the clinical profile with causes of death in deceased patients admitted in a COVID-19 dedicated hospital in Dhaka, Bangladesh.

**Methods:** This cross-sectional retrospective study included 108 COVID-19 associated deceased patients admitted in Kurmitola general hospital, Dhaka, Bangladesh between 25 March 2020 and 24 June 2020) Data were collected from hospital record. Causes of death were categorized into early and late with cut-off of 48 hours of hospitalization.

**Results:** Among 809 hospitalized cases of COVID-19, 108 patient died (13.35%) over three months of study period. The mean age of the deceased patients was 60.22±13.94 years; 86.1% were male. About 85% had at least one comorbidity with diabetes mellitus (65.7%) was the most common one. The most common symptoms were breathlessness (88.0%), fever (65.7%) and cough (43.5%). Nearly 75% presented with severe disease. Patients had altered biochemical profiles and treated with different drugs including antibiotics and steroids. Young age and malnutrition were two characteristic features. Only one third got intensive care support. The most common cause of death was acute respiratory syndrome (95.37%). Septic shock & acute myocardial infarction were predominantly early and uremia, hepatic failure & hyperglycemic crisis were the predominant causes of late hospital death.

**Conclusions:** The findings of this study will help clinicians as well as policy makers to take necessary steps to prevent death from COVID-19 in Bangladeshi population.

## Introduction

The coronavirus disease 2019 (COVID-19), caused by severe acute respiratory distress syndrome virus 2 (SARS-CoV-2) has infected more than 18 crore people along with around four million deaths worldwide as of 04 July 2021. In Bangladesh, over 14,000 deaths have been reported due to COVID-19. Currently, the situation is expected to become worse, perhaps because of new delta variant. The majority of the patients with COVID-19 recover but only a small portion of the affected persons are at increased risk of fatal outcome^1^. Most of the studies reported several co- morbidities as risk factors of COVID-19 death.^2^ However, autopsy reports suggest that COVID- 19 is the direct cause of death with little contribution from preexisting health conditions. Septic shock, multi-organ failure, acute and organizing diffuse alveolar damage as well as pulmonary thromboembolism etc. are reported as immediate causes of death in COVID-19 patients.^3,4^ The characteristics of deceased person as well as causes of death may help clinicians to identify population at risk and take appropriate management. However, the information on this aspect of COVID-19 deaths is lacking in South Asian countries, particularly from Bangladesh. The aim of this study is to describe the clinical and laboratory characteristics along with causes of death in patients with COVID-19.

## Materials and Methods

This retrospective cross-sectional study was conducted among 108 deceased patients of RT-PCR confirmed COVID-19 who had been admitted in Kurmitola General Hospital (KGH), Dhaka during the period of 25 March, 2020 to 24 June, 2020. Data were extracted from hospital records using a relevant questionnaire. All investigation findings done in the 1^st^ time were included. All information were double-checked before analysis to ensure quality. The severity of COVID-19 was described by WHO interim guidance and the period of death was divided in to early and late by cut-off of 48 hours of hospitalization.^5,6^ The institutional review board of the Biomedical Research Foundation, Bangladesh, approved the study protocol (Ref. no: BRF/ERB/2020/003). Data were expressed in mean± standard deviation (SD) or median (interquartile range, IQR) or number (percentages, %). Analysis was done among available data for each variable.

## Results

Among 809 hospitalized cases of COVID-19, 108 patient died (13.35%) over three months of study period. The baseline characteristics of deceased patients with COVID-19 are shown in Table 1. The median age of the study population was 60.0 (50.0, 70.0) years (min-max: 30 - 99 years) with 49% of the patients were above 60 years. Most of the patients were male (86.1%) and had residence in urban area (80.6%). Only 15.7% patients had no comorbidity while around 60% of them had at least two comorbidities. The most common comorbidities were diabetes mellitus (DM) (65.7%) followed by hypertension, chronic kidney disease (CKD), ischemic heart disease (IHD) etc. More than half of the patients died within 4 days of admission

**Table 1.** Baseline characteristics of deceased COVID-19 patients (N= 108)

| **Variables** | **Study population** |
| --- | --- |
| Number | 108 |
| Age (years), median (IQR) | 60.0 (50.0, 70.0) |
| Age groups, no. (%) |  |
| ≤40 years | 11 (10.2) |
| 41 – 60 years | 44 (40.7) |
| 61 – 80 years | 44 (40.7) |
| ≥80 years | 9 (8.3) |
| Sex, no. (%) |  |
| Male | 93 (86.1) |
| Female | 15 (13.9) |
| Residence, no. (%) |  |
| Urban | 87 (80.6) |
| Rural | 21 (19.4) |
| Blood group, no. (%) | [26]* |
| A | 11 (42.3) |
| B | 9 (34.6) |
| O | 5 (19.2) |
| AB | 1 (3.8) |
| Number of comorbidities, no. (%) |  |
| 0 | 17 (15.7) |
| 1 | 29 (26.9) |
| ≥2 | 62 (57.4) |
| Comorbidities, no. (%) |  |
| DM | 71 (65.7) |
| Hypertension | 51 (47.2) |
| CKD | 31 (28.7) |
| IHD | 21 (19.4) |
| Bronchial asthma | 10 (9.3) |
| CVD | 5 (4.6) |
| CLD | 3 (2.8) |
| COPD | 2 (1.9) |
| Malignancy | 2 (1.9) |
| Hospital stay, days | 5.0 (2.0, 8.0) |
| 0 – 4 | 56 (51.9) |
| 5 – 9 | 33 (30.6) |
| ≥ 10 | 19 (17.6) |

Pregnant (1), hypothyroid (3), BEP (1), parkinson's disease (2), etc; \*[available number]

The average days of onset of symptoms to hospital admission was more than 8 days (8.17±3.08). The most common presenting complaints were breathlessness (88%), followed by fever (65.7%) and cough (43.5%). Less common presenting symptoms were vomiting, runny nose, bleeding, altered consciousness, anorexia, sore throat, chest pain, abdominal pain, fatigue. Four patients died without any documented symptoms. While tachycardia was present in 57.4%, pyrexia was present in only one patient at presentation. Oxygen saturation was critically low in 70.4% and 74.1% presented with severe and critical status (Table 2).

**Table 2.** Clinical features of deceased patients with COVID-19 at presentation (N= 108)

| Variables | Study population |
| --- | --- |
| Symptoms at presentation |  |
| Onset to hospitalization (days), mean±SD | 8.17±3.08 |
| Asymptomatic, no (%) | 4 (3.7) |
| Difficulty in breathing, no (%) | 95 (88.0) |
| Fever, no (%) | 71 (65.7) |
| Cough, no (%) | 47 (43.5) |
| Signs at presentation |  |
| Pulse (beats/ minute), mean±SD | 104.23±23.40 |
| Tachycardia (pulse >100 beats/minute) | 62 (57.4) |
| Temperature ($^{\circ}$C), mean±SD | 36.0±0.64 |
| Pyrexia (Temperature >38 $^{\circ}$C) | 01 (0.9%) |
| Oxygen saturation (%), mean±SD | 79.19±16.03 |
| Low oxygen saturation (<90%) | 76 (70.4) |
| Severity of disease at presentation |  |
| Nonsevere (mild to moderate), no (%) | 28 (25.9) |
| Severe (severe to critical), no (%) | 80 (74.1) |

Other symptoms: Vomiting (4), runny nose (4), bleeding (3), altered consciousness (3), anorexia (2), sore throat (2), chest pain (2), abdominal pain (1), fatigue (1), etc.

Among 49 available complete blood count reports, more than half had anemia. While leukocytosis was present in 77.6%, both lymphopenia and thrombocytopenia were present in 22.4% of patients. High neutrophils/lymphocytes ratio and platelets/lymphocytes ratio were present in 73.5% and 36.7% of patients respectively. Among 74 available electrolyte reports, nearly 50% patients had hyponatremia. Hyperkalemia (27%) was more frequent than hypokalemia (10.8%). The mean/median values of all the biochemical variables were abnormal than their respective reference range and a significant percentages of the study population had different abnormal values (Table 3).

**Table 3.**
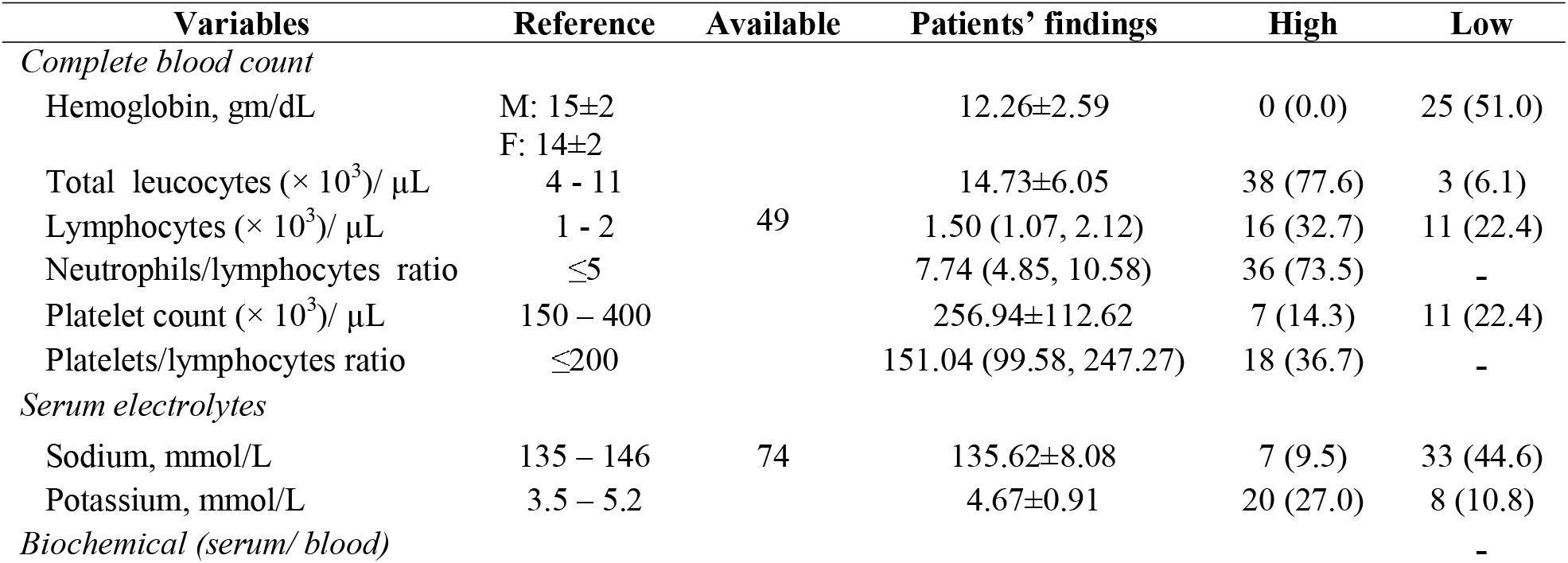
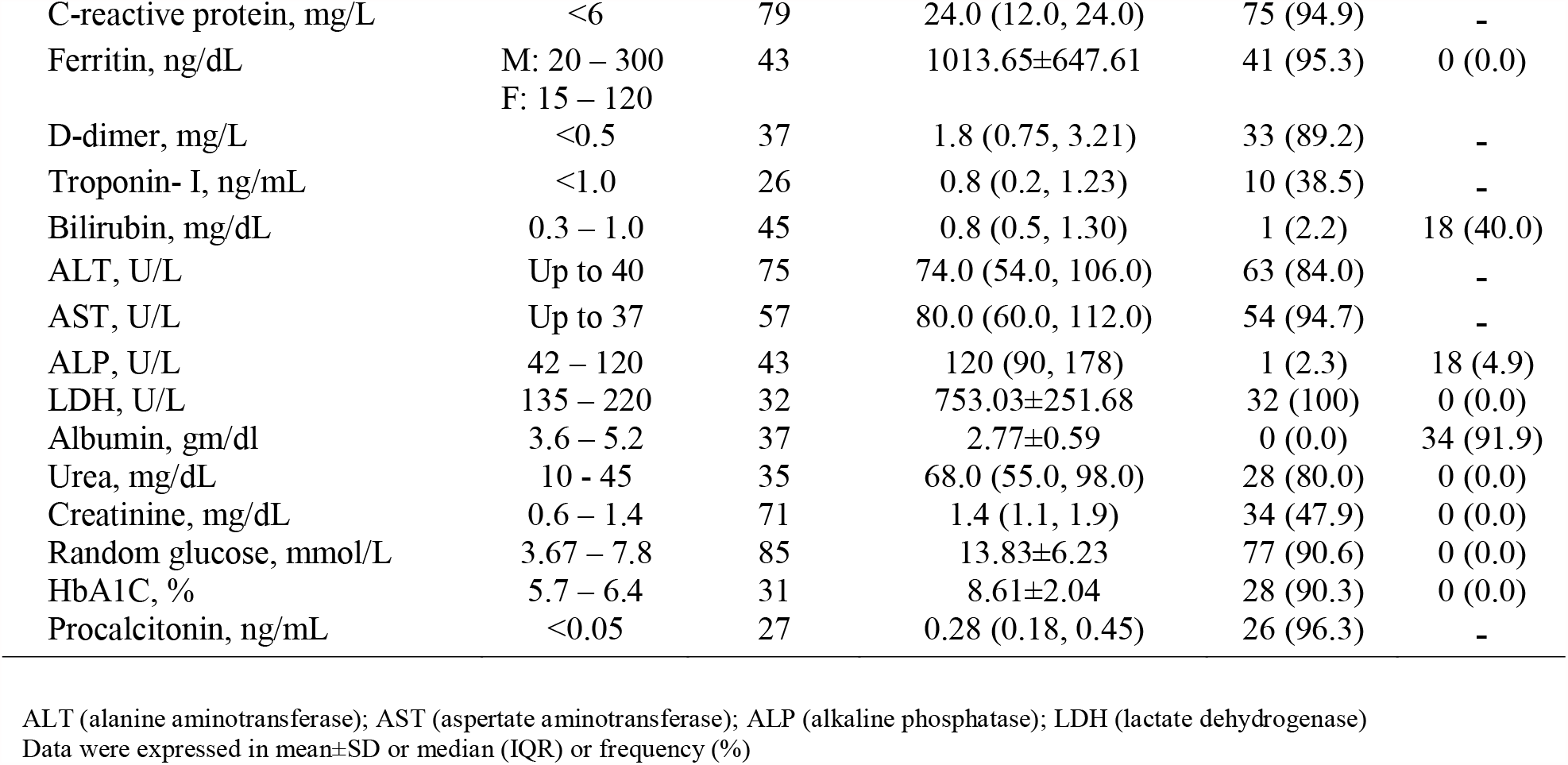
Investigation findings in deceased patients with COVID-19.

While different types of antibiotics both in oral and parenteral routes were prescribed in around two thirds of patients, antivirals were used in around one third of cases. A few cases were treated with ivermectin, hydroxychloroquine, tocilizumab and convalescent plasma therapy. Around 60% patients received different types of steroids. Insulin was more prescribed than oral antidiabetic agents. (Table 4).

**Table 4.**
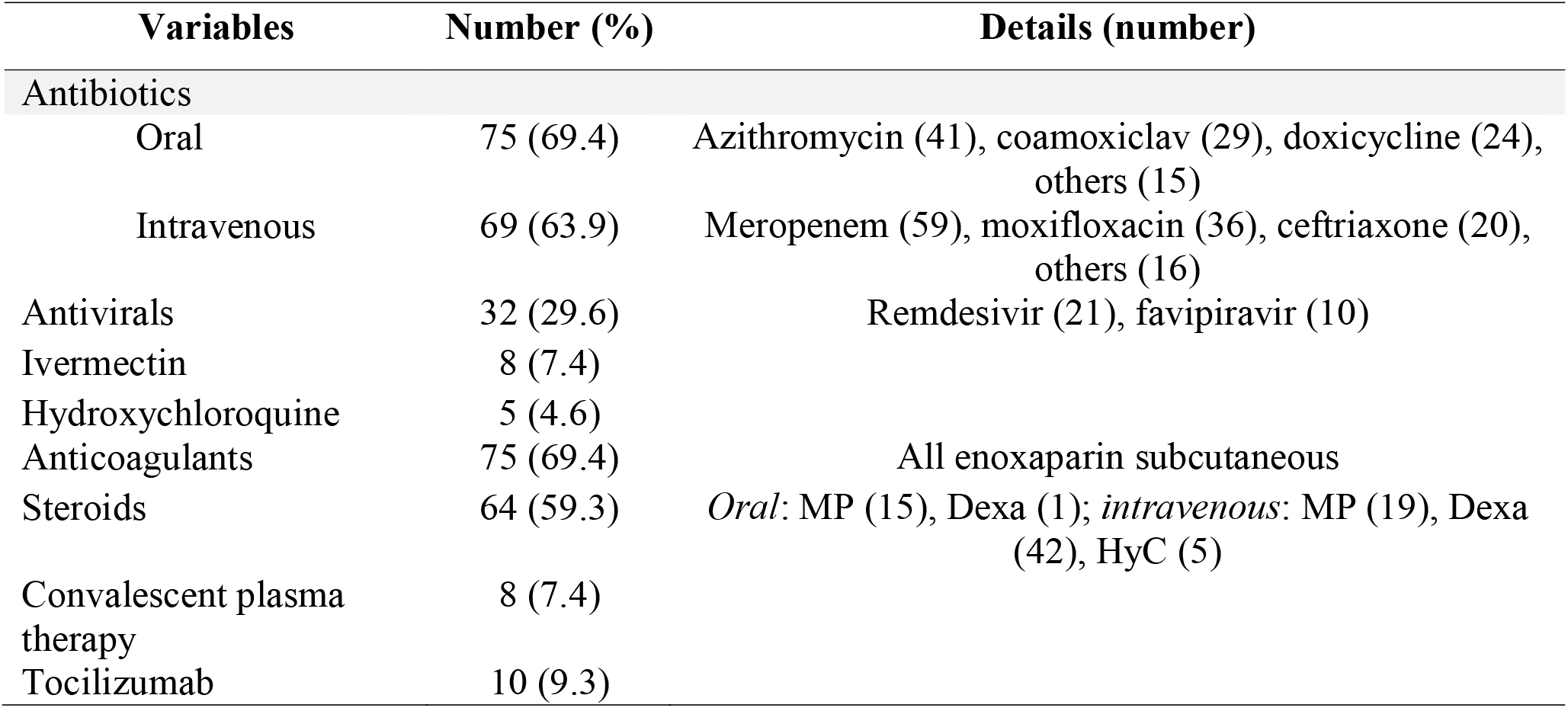
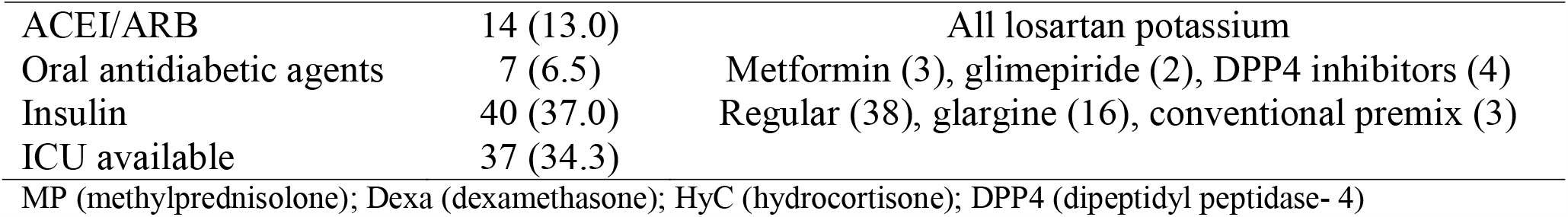
Treatment and cause of death of deceased COVID-19 patients.

Figure 1 is showing the causes of deaths in patients with COVID-19 according to duration of hospital stay (cut-off of 48 hours). The most common cause of death was acute respiratory distress syndrome (95.3%) which occurred at a same rate in early and late period of hospitalization. While septic shock and acute myocardial infarction were the predominant causes of early death, uremia was more common among patients with late deaths. Hepatic failure, hyperglycemic crisis and hemorrhage were exclusively late causes of death. Electrolyte imbalance was another cause of death which has affected both the hospital periods.

**Figure 1.**
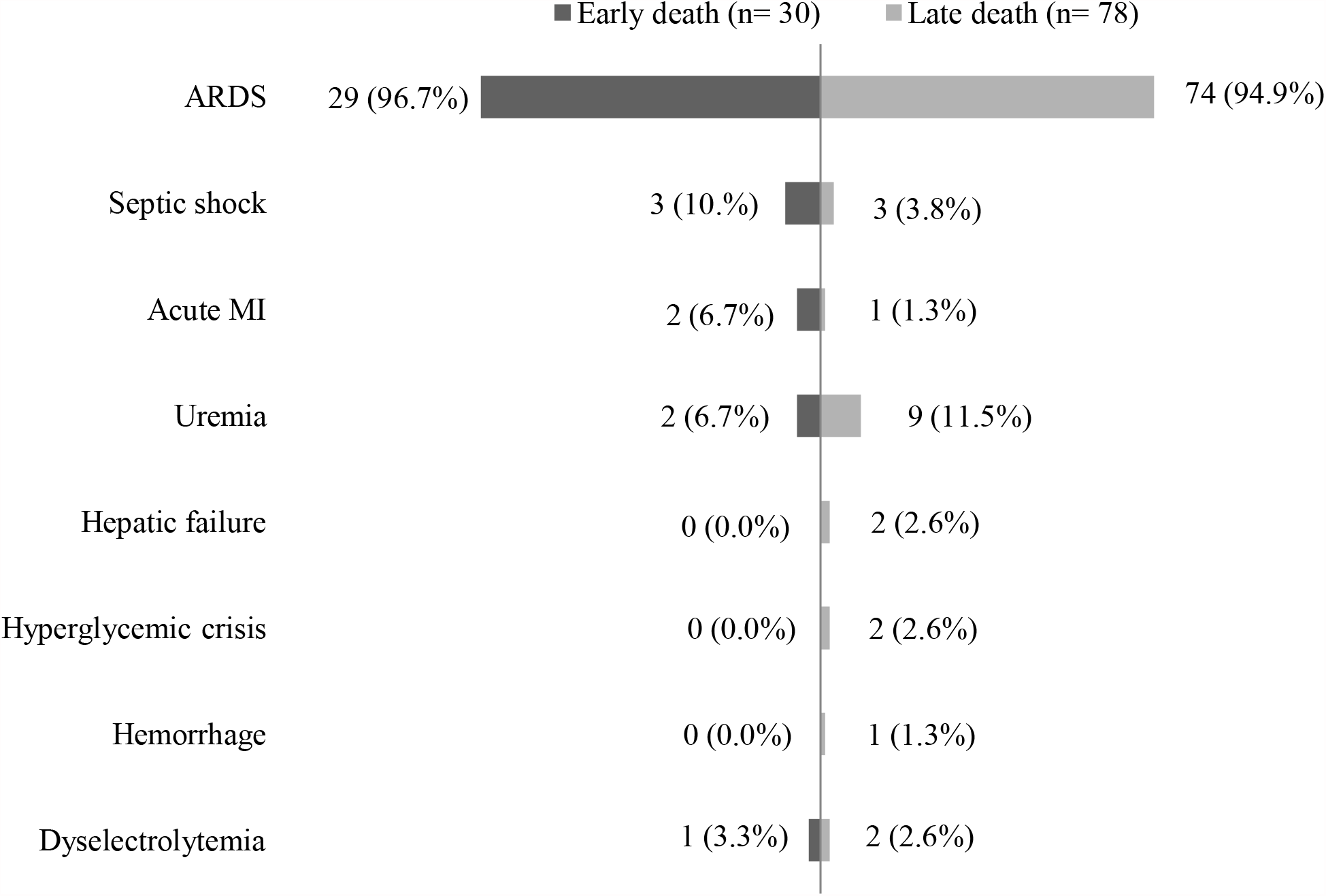
Causes of death with duration of admission (cut-off of 48 hours)

## Discussion

This study has revealed a death rate of 13.3% during the first three months from initiation of hospitalization of COVID-19 patients. Our finding is consistent with studies reported from Asian countries.^2^ Age ≥65 years was found as a major risk factor for death in patients with COVID-19 in a systematic review and meta-analysis of observational studies.^7^ Interestingly, more than half of our study population were younger than 60 years. This finding is in concordance with recent reviews from Bangladesh and India;^8,9^ suggesting that population at risk for COVID-19 mortality may be different from other regions of the world possibly due to genetic or environmental factors or possibly a shorter life expectancy. Other demographic characteristics, clinical features, co-morbidities and duration of hospital stay were similar to other studies.^10-12^

More than half of the study population had anemia with low albumin level indicating under nutrition as a probable contributing factor of death in COVID-19 patients. Previous studies highlighted obesity, however, the scenario may be different in a developing country like Bangladesh which faces the double burden of malnutrition.^13^ However, data regarding body mass index was not documented in hospital records.

More than 90% of deceased patients had high neutrophils/lymphocytes ratio, platelets/lymphocytes ratio, c-reactive protein, ferritin and procalcitonin indicating a severe inflammatory response. Similarly, a significant proportion of patients with high D dimer, suggesting a prothrombotic state in COVID-19.^14-16^ High liver enzymes and creatinine were probably systemic responses of COVID-19 infection and may affect prognosis.^17^ High random glucose in around 90% of patients were due to inflammatory response along with steroid use. High HbA1C, found among patients with diabetes mellitus also indicates poor glycemic control, an important risk factor for poor outcome.^18^

Several treatment options were tried initially without much evidence. However, most patients could not able to manage ICU due to unavailability in 10-bedded ICU in the study hospital. We found ARDS as the main cause of death in patients with COVID-19 rather than associated co- morbidities. Slater et al. (2020) also showed death in COVID-19 as a direct consequence of the virus similar to an autopsy report of 26 deceased COVID-19 patients.^3,19^

There are several limitations of our study. The sample size is small. All the data are retrospective and secondary with a possibility of under-reporting along with many incomplete data.

## Conclusions

Our study findings showed several characteristics and unique features (young age, under nutrition) of deceased patients with COVID-19 than other parts of the world. These findings would help clinicians and policy makers to tailor management strategies, facilitate decision making and ultimately improve patient outcome.

## Data Availability

All available data has been included

## Acknowledgments

We are grateful to Brigadier General Jamil Ahmad, director, Kurmitola General Hospital (KGH); Md. Ashek Ali, statistical officer, KGH for their generous support in data collection.

## Declarations

### Ethics approval and consent to participate and publish

The institutional review board of Biomedical Research Foundation, Bangladesh approved the study protocol (Ref. no: BRF/ERB/2020/003).

